# Personalized Risk Stratification in Colon Cancer using Radiomic-Based Predictive Models

**DOI:** 10.1101/2025.11.27.25341095

**Authors:** Manuel Collado, María E. Castillo, María Jesús Larriba, Cristina Galindo-Pumariño, Carmen-Lisset Flores, Reyes Ferreiro, Elena Canales, Raquel García, José Avendaño-Ortiz, José Manuel Gonzalez-Sancho, Carolina de la Pinta, Cristina Peña

## Abstract

Colon Cancer (CC) is among the most frequently diagnosed malignancies and a leading cause of cancer-related death worldwide. Five-year survival varies markedly by stage at diagnosis, underscoring the need for precise risk stratification. Otherwise, although stage III CC patients routinely receive adjuvant chemotherapy, some exhibit a low risk of recurrence and could safely undergo shorter regimens. Conversely, a subset of stage II patients faces a higher relapse risk and may benefit from intensified treatment. In this context, radiomics has emerged as a cutting-edge, non-invasive approach capable of extracting quantitative information from routine medical imaging to support clinical decision-making. We aimed to develop radiomic-based machine learning models able to distinguish stage II from stage III CC patients while identifying individuals at increased risk of 5-year relapse. A cohort of 104 patients who underwent preoperative computed tomography was analyzed by 3D tumor segmentation. Thus, 105 radiomic features and preoperative clinical variables were extracted. Predictive models were trained and validated employing a 70/30 split and 10-fold cross-validation. The Generalized Linear Model achieved the best performance for stage differentiation (AUC=0.760). For 5-year relapse prediction, the Partial Least Squares model showed excellent performance (AUC=0.910), outperforming the Support Vector Machine model (AUC=0.730). Subgroup analyses confirmed strong predictive capacity when evaluating stage II (AUC=0.929) and stage III patients (AUC=0.926) separately. To our knowledge, this is among the first studies demonstrating that radiomics can simultaneously stratify CC stage and predict relapse risk with high accuracy, highlighting its potential as a powerful tool to guide personalized treatment strategies.

**Graphical abstract:** 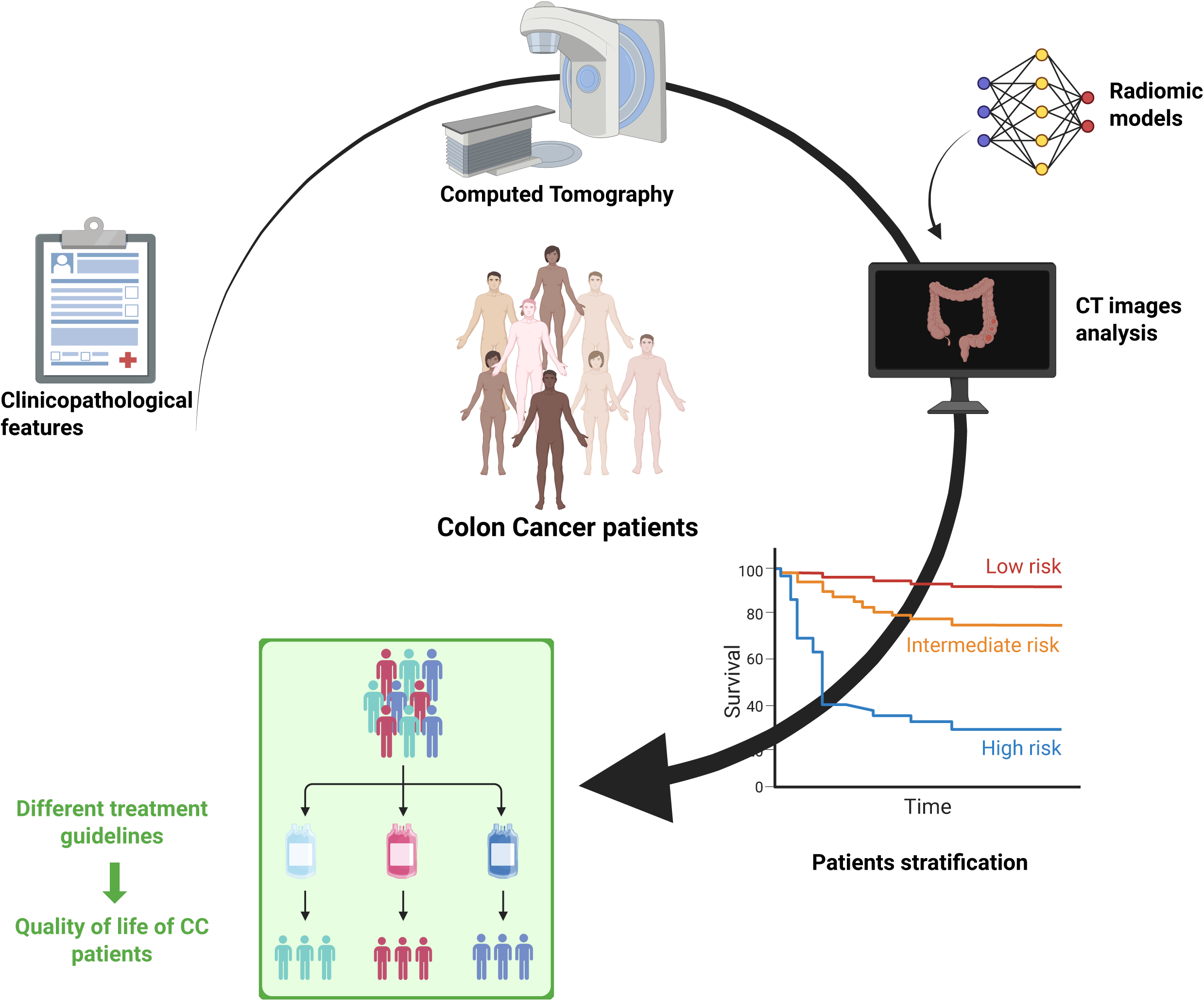

## 1. INTRODUCTION

### 1.1. Clinical background and unmet needs

Colon cancer (CC) is among the most commonly diagnosed cancers and a leading cause of cancer-related mortality worldwide (Filho et al., 2025). Adjuvant chemotherapy clearly improves outcome in stage III CC, increasing the 5-year disease-free survival (DFS) rate from 49.0% to 63.6% (Böckelman et al., 2015). However, approximately 25–30% of stage III CC patients experience relapse within five years due to minimal residual disease that remains after surgery and chemotherapy (Brenner et al., 2014; Murray et al., 2019). Conversely, patients with a low risk of recurrence could benefit from shorter chemotherapy regimens, thereby avoiding unnecessary toxicity (Grothey et al., 2018). Improving risk stratification is therefore critical for individualized patient management.

The tumor–node–metastasis (TNM) staging system proposed by the American Joint Committee on Cancer (AJCC) is the standard classification for patient management. However, the heterogeneous outcomes observed among patients with the same tumor stage question its usefulness at an individual level (Amin et al., 2017). This variability is mainly attributed to tumor heterogeneity, which contributes to phenotypic and functional diversity among cancer cells and may affect treatment sensitivity (Shackleton et al., 2009). Several clinicopathological markers, such as deficient mismatch repair (dMMR), microsatellite instability-high (MSI-H) status (Diaz et al., 2022), or BRAF^V600E^ mutation (Tabernero et al., 2021), have been proposed as prognostic indicators in CC, yet their predictive performance remains suboptimal (Fabregas et al., 2022). Consequently, the development of new predictive models and more accurate prognostic biomarkers is crucial to advance toward individualized patient management and personalized medicine.

### 1.2. Role and limitations of current imaging in Colon Cancer management

In current clinical practice, the diagnostic and staging work-up of CC typically involves colonoscopy followed by biopsy, which is invasive and may fail to represent tumor heterogeneity. These procedures are complemented by laboratory testing and imaging studies, most notably computed tomography (CT) (Argilés et al., 2020). Preoperative CT is mainly used to exclude synchronous metastases, but its prognostic potential remains insufficient (Smith et al., 2007). However, emerging evidence suggests that CT imaging may also provide valuable information related to future disease progression and treatment response (Lee et al., 2023).

### 1.3. Study objectives and contributions

Using clinical and imaging data, this retrospective study sought to stratify CC patients into stage II and stage III and to predict 5-year relapse through radiomic features derived from preoperative CT scans. As a non-invasive and readily available imaging-based approach, radiomics provides prognostic information even before surgery, offering a unique opportunity to refine risk assessment at an early stage of patient management. The insights gained from this work could enable more accurate treatment selection, paving the way for personalized therapeutic strategies and ultimately improving outcomes and quality of life in CC patients.

## 2. PRIOR WORK

### 2.1. Potential of radiomics to aid in clinical decision-making

In recent years, the analysis of diagnostic imaging techniques has gained increasing relevance in oncology. Radiomics, a non-invasive quantitative approach in medical imaging, goes beyond visual interpretation by extracting a large number of mathematical features that characterize tumor morphology and texture. These features can be integrated with clinical data to develop decision-support machine learning models based on computational algorithms (Gillies et al., 2016; Mayerhoefer et al., 2020; Yip & Aerts, 2016). Radiomic features capture properties such as concavity, irregularity, or sharpness of tissue morphology, as well as texture descriptors like asymmetry, kurtosis, and entropy, providing a deeper characterization of lesion microarchitecture (Aerts, 2016).

Radiomics has already shown promising results in several solid tumors. It has been used to predict survival in non-small cell lung cancer patients with brain metastases when combined with RNA sequencing data (Deng et al., 2025), and to predict immunotherapy response in conjunction with clinical variables (He et al., 2020; Liao et al., 2024). Radiomic features have successfully distinguished breast cancer molecular subtypes (Hu et al., 2024; Yang et al., 2025; M. Yao et al., 2025) and assessed axillary lymph node status in patients receiving neoadjuvant therapy (J. Yao et al., 2024; Yu et al., 2024)., It has also been applied to predict response to neoadjuvant chemoimmunotherapy in head and neck cancer (Lin et al., 2024) and to detect adverse effects of radiotherapy, such as mandibular osteoradionecrosis (Kamel et al., 2025) or xerostomia (Khajetash et al., 2025). Moreover, radiomic models have shown potential for determining MGMT methylation status (Chen et al., 2025) and IDH1 mutation in gliomas (Liang et al., 2024; Ye et al., 2025).

### 2.2. Lack of true clinical utility of radiomics in Colon Cancer

In colon cancer, however, published studies remain relatively limited and report heterogeneous results. Several studies have investigated the potential of CT-derived radiomic biomarkers to predict prognosis in CC (Caruso et al., 2022; Dai et al., 2020; Li et al., 2019), and some have explored associations with histopathological parameters or mutational profiles. For example, Negreros-Osuna *et al*. demonstrated the utility of CT-based radiomic biomarkers for identifying BRAF mutation and predicting 5-year overall survival (OS) in stage IV CC patients (Negreros-Osuna et al., 2020). Similarly, other studies have explored associations between texture features and KRAS mutational status in colorectal cancer liver metastases (MG et al., 2015). Despite these promising findings, the variability in imaging protocols, feature extraction pipelines and study designs has made it difficult to establish the true clinical potential of radiomic in this disease.

## 3. MATERIALS AND METHODS

### 3.1. Colon Cancer Patients

This study was approved by the Ethics Committee of Ramón y Cajal University Hospital. The initial database included 121 CC patients recruited at the same institution, all of whom provided written informed consent to participate in the study. All patients underwent colonoscopy, preoperative CT, surgery, and adjuvant chemotherapy when indicated. Preoperative CT images and postoperative pathological reports were collected for all cases.

After applying the inclusion and exclusion criteria, the final study cohort comprised 104 CC patients. They were randomly divided into training (70%) and validation (30%) cohorts, ensuring a comparable distribution of imaging and clinical features across both sets.

Clinical data—including sex, age at diagnosis, primary tumor location, histopathological features, and tumor stage (according to the TNM staging system)—were collected from the hospital information system.

### 3.2. Computed Tomography Image Acquisition and Feature Extraction

CT scans were performed between December 2016 and September 2022 and uploaded to the QP-Precision platform, which was specifically deployed for this study by Quibim company. Using this platform, the primary tumor was manually segmented in three dimensions (3D), slice by slice, by trained image technicians from Quibim in collaboration with radiology experts from Ramón y Cajal University Hospital. Figure 1A illustrates the segmentation of the primary lesion in one representative case (patient 82).

**Figure 1.**
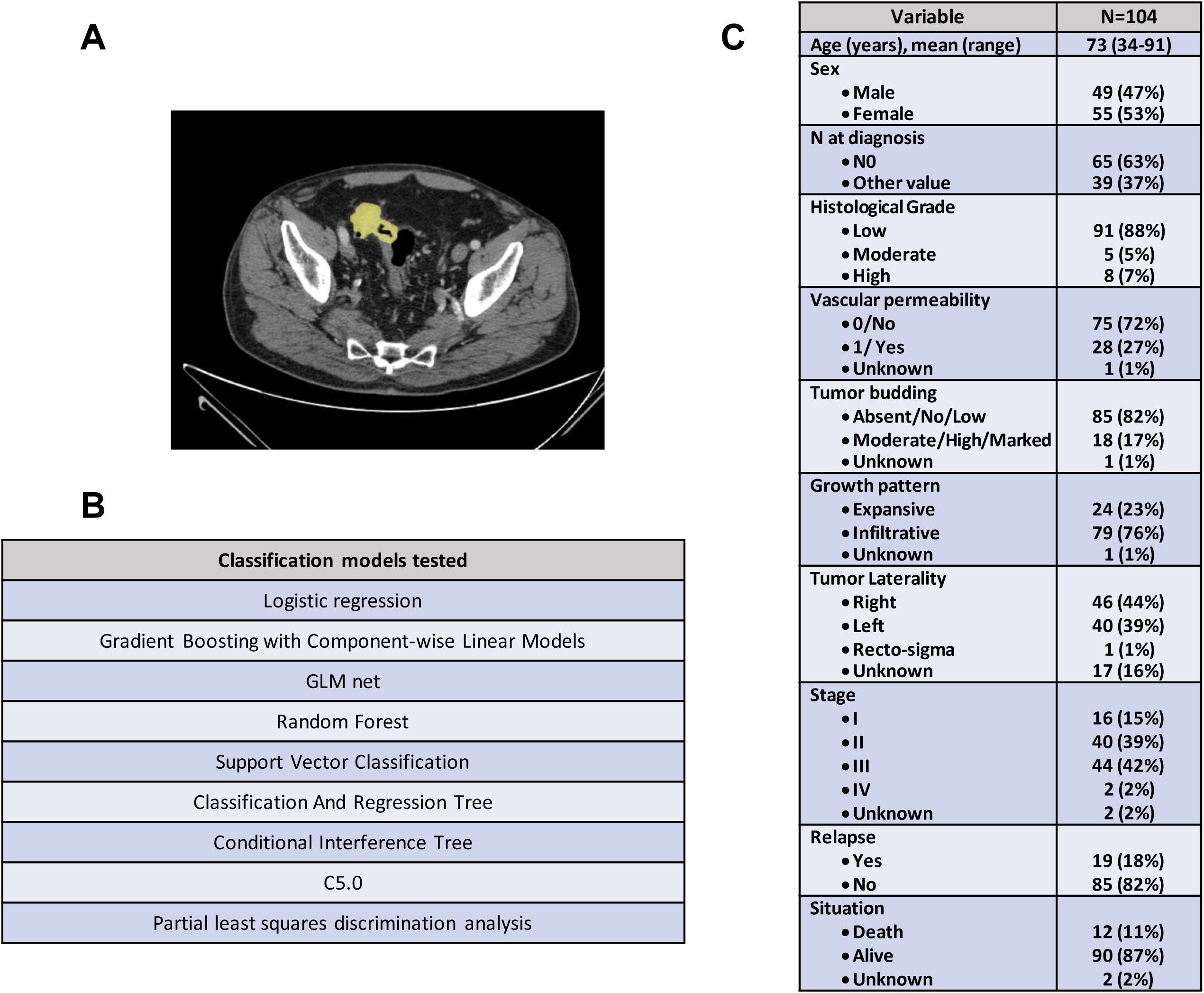
Clinical and imaging data of the cohort of colon cancer patients. A) In yellow segmentation of CT images of the primary tumor of patient 82 using Quibim Precision® platform. B) Classification models tested in this study. C) Clinicopathological characteristics of the patient cohort.

Texture analysis of the segmented CT lesions was performed for all 104 cases. These imaging biomarkers provide quantitative information related to tissue heterogeneity, which is often associated with tumor aggressiveness and progression. From each region of interest (ROI), a total of 105 quantitative radiomic variables were extracted and categorized (Scapicchio et al., 2021) into the following groups:

- **Shape features:** Quantitative descriptors of the geometric characteristics of the ROI, including surface area, volume, maximum diameter, elongation, sphericity, and surface-to-volume ratio.
- **First-order statistics (histogram-based features):** Metrics describing the distribution of voxel intensities within the ROI, including energy, entropy, mean, interquartile range, skewness, kurtosis, and uniformity.
- **Second-order statistics (textural features):** Parameters derived from matrices that capture spatial relationships between neighboring voxels including:

**- Gray-Level Co-occurrence Matrix (GLCM):** Describes spatial relationships and gray-level intensity distributions within the 3D volume.
**- Gray-Level Run-Length Matrix (GLRLM):** Measures the number of consecutive voxels sharing the same gray-level value, providing information about run lengths across different directions.
**- Gray-Level Size-Zone Matrix (GLSZM):** Quantifies homogeneous zones defined as clusters of connected voxels with identical intensity values.
**- Neighboring Gray-Tone Difference Matrix (NGTDM):** Captures contrast between each voxel intensity and the mean intensity of its neighboring voxels within a specified distance (δ).
**- Gray-Level Dependence Matrix (GLDM):** Assesses the number of connected voxels within a distance δ that depend on the intensity of a central voxel.

A descriptive analysis of the radiomic variables was performed to identify anomalous data and near-zero variance features. This process is detailed in Supplementary Material I.

### 3.3. Predictive Models

Multiple predictive machine learning modeling techniques were compared to select the optimal model for each study endpoint based on their statistical performance. The primary endpoints of the study were:

- Differentiation between stage II and stage III CC patients. For this analysis, the cohort was reduced to 84 patients after excluding those with stage I, IV, or unknown.
- Prediction of 5-year relapse from diagnosis, including the full cohort (N = 104).

For both endpoints, radiomic features (105 variables per ROI) were combined with 9 clinical and pathological variables: age, sex, tumor laterality (left/right), T stage (<4 vs. >4), N stage (N0 vs. N+), histological grade (low vs. moderate/high), vascular permeability (yes/no), growth pattern (infiltrative vs. expansive) and tumor budding (none/low vs. moderate/high).

The dataset was split into a training set (70%) and a validation set (30%). A 10-fold cross-validation strategy was applied within the training set to prevent overfitting. Therefore, the datasets used were:

- Stage II vs. III differentiation: total N = 84 (training N = 59; validation N = 25)
- 5-year relapse prediction: total N = 104 (training N = 73; validation N = 31)

#### 3.3.1. Imputation of Missing Variables

For variables with missing data, imputation was performed using the k-Nearest Neighbors (KNN) algorithm, which assigns missing values based on the nearest neighbors within the dataset. For the first endpoint (stage differentiation), tumor laterality was missing in 15 patients, and one patient lacked data for the T parameter and perineural invasion. For the second endpoint (5-year relapse), missing data were: laterality (17), T (1), vascular invasion (1), perineural invasion (2), lymphatic invasion (2), growth pattern (1), tumor budding (1), and peritumoral lymphocyte response (1).

#### 3.3.2. Dimensionality Reduction and Classification Models

Radiomic variables with near-zero variance (<0.01) were excluded from the analysis. According to this criterion, 45 variables were removed, resulting in a final set of 63 quantitative features. The classification models evaluated for each of the study objectives are summarized in Figure 1B.

## 4. RESULTS

### 4.1. Clinicopathological characteristics of the study cohort

The final study cohort included 104 patients diagnosed with CC with a mean age of 73 years (range: 34-91) and comprising 49 males and 55 females. Tumor location was right-sided in 46 (44%) patients and left-sided in 40 (39%), while 1 tumor was located in the rectosigmoid junction (1%) and 17 had an unknown location (16%). Regarding tumor stage, 16 patients (15%) were classified as stage I, 40 as stage II (39%), 44 as stage III (42%), 2 as stage IV (2%), and 2 with unknown stage (2%). During the follow-up period, 19 patients experienced relapse (Figure 1C).

### 4.2. Patient stratification by stage (II vs. III)

As expected, when combining the 9 clinical and pathological variables with the radiomic features, pathological variables, particularly the N parameter, showed a strong discriminative ability to separate stage II from stage III patients (data not shown). However, since the main objective of this study was to identify new imaging biomarkers that could be available prior to surgery, it was not appropriate to include pathological variables in this endpoint. Therefore, they were excluded, and only three preoperative clinical variables—age, sex, and tumor laterality (left/right)—were included in the study.

As described in the Methodology section, missing data were imputed using the KNN model. Variables showing near-zero variance (<0.01) were removed, and a univariate analysis using the Mann–Whitney U test was conducted. Only variables with an effect size >0.10 and p <0.05 were retained. After this process, four radiomic features were selected for the classification model (Figure 2A). Detailed results, including p-values and effect sizes for all variables, are provided in Supplementary Material II.

**Figure 2.**
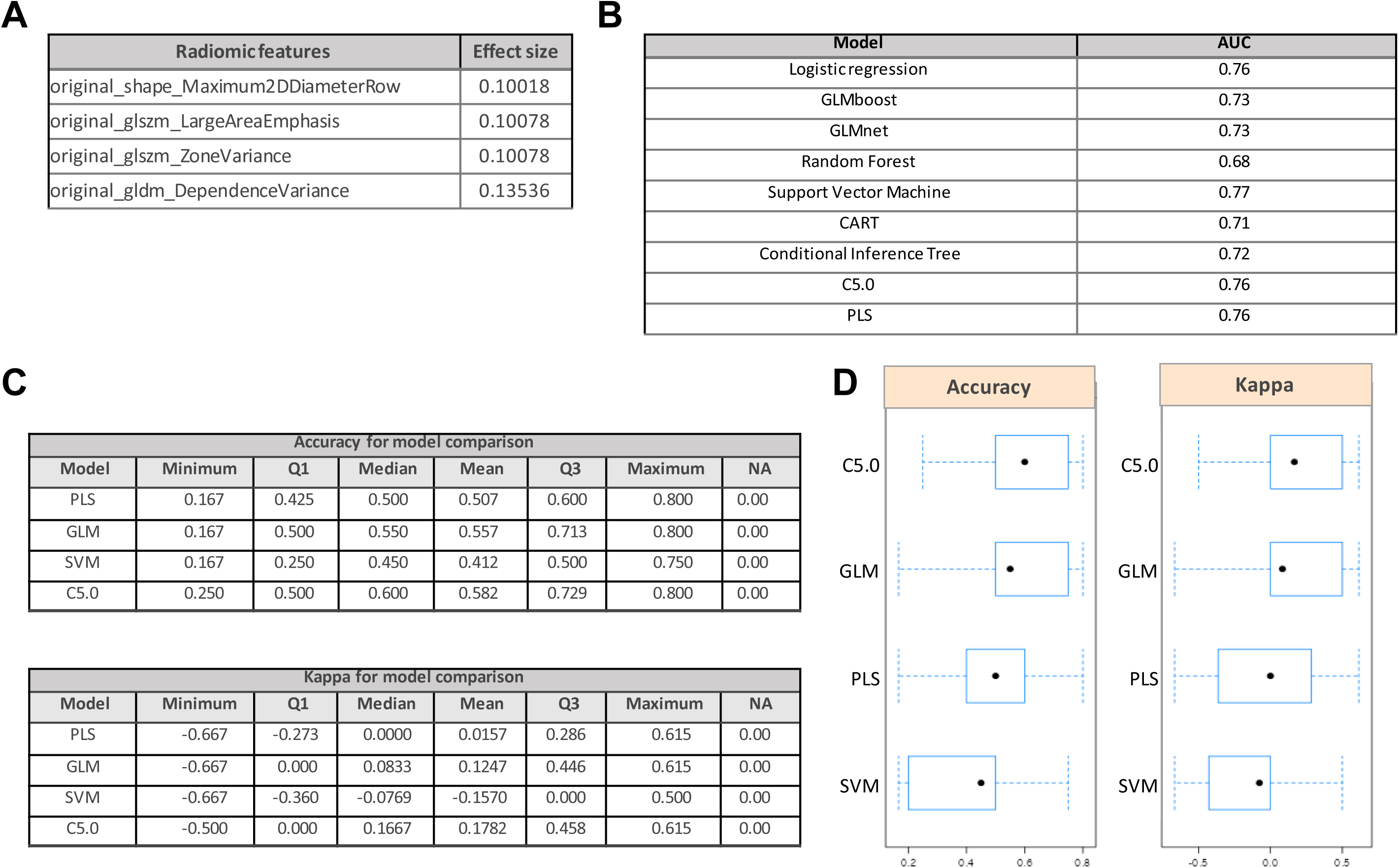
Radiomic data and model performance for the stratification of stage II and stage III colon cancer patients. A) Radiomic features included in the patient stratification analysis. B) AUC values of the different classification models tested for stage II vs. stage III stratification. C) Comparison of model performance using accuracy and kappa metrics for the endpoint of stage II vs. stage III stratification. D) Boxplots comparing classification models based on accuracy and kappa metrics for the endpoint of stage II vs. stage III stratification.

The dataset was randomly split into 70% for training and 30% for testing, with model parameters optimized through 10-fold cross-validation. Multiple classification models were evaluated, most of which performed well, achieving an Area Under the Curve (AUC) above 0.7 (Figure 2B). However, to identify the model with the best performance, we compared them using two evaluation metrics: accuracy and kappa (Figure 2C and 2D).

Although the C5.0 model showed slightly higher accuracy, the Generalized Linear Model (GLM) was ultimately selected due to its interpretability, statistical inference capability, and overall fit. The GLM achieved a sensitivity of 0.889 for predicting stage II patients with a balanced accuracy of 0.744 (Figure 3A). The relative importance of the included variables is shown in Figure 3B. The ROC curve for the GLM model yielded an AUC of 0.760 (Figure 3C).

**Figure 3.**
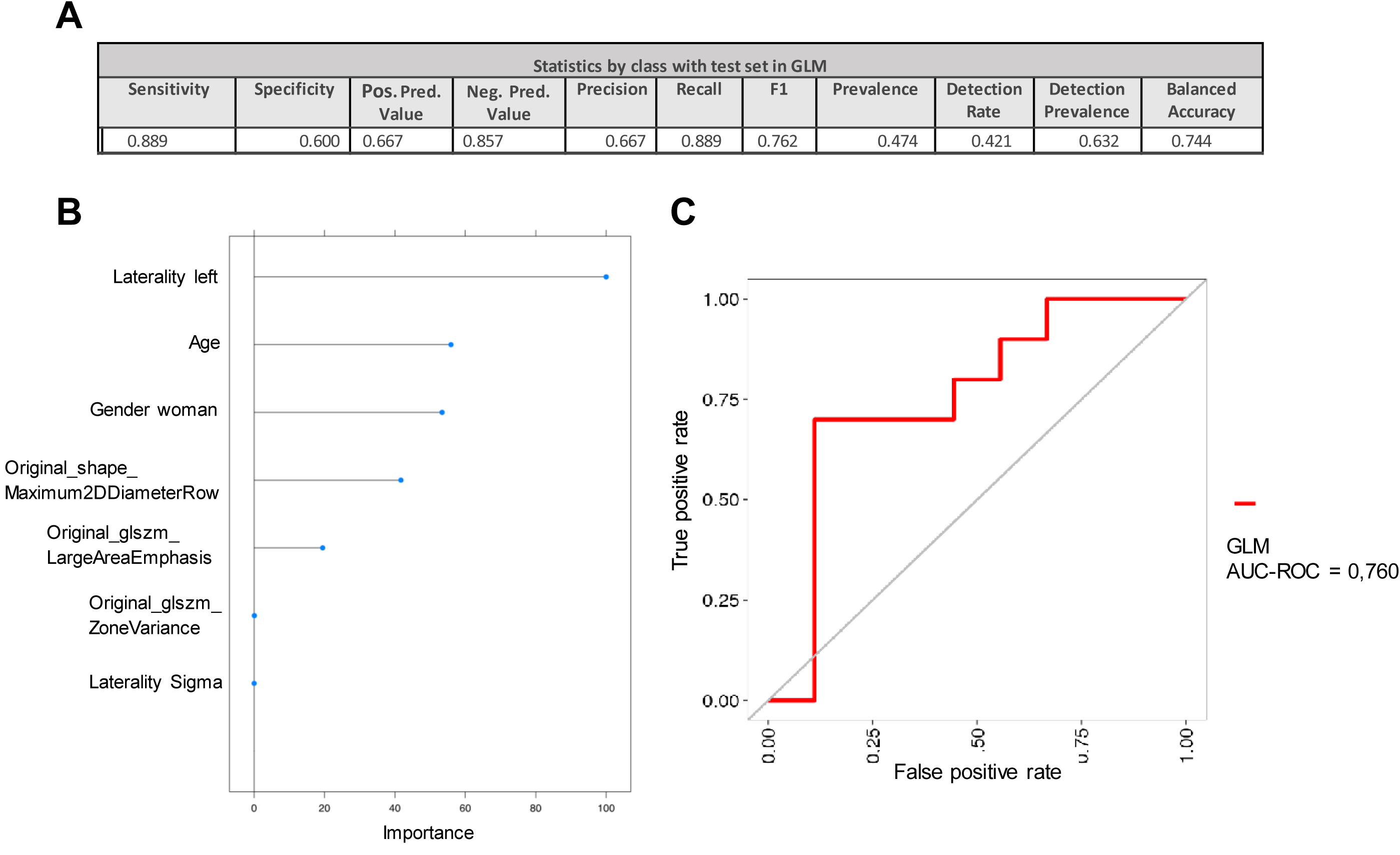
Preoperative radiomic data stratify colon cancer patients into stage II and stage III. A) Class statistics for the test set using the GLM model for patient stratification. B) Variable importance in the GLM model for patient stratification. C) ROC curve for the GLM model in the stratification between stages II and III.

### 4.3. Prediction of 5-year relapse in colon cancer patients

To develop a model for predicting 5-year relapse, a similar approach, including preoperative clinical variables, was applied. Missing data were imputed with the KNN model, and variables with low variance (<0.01) were excluded. A univariate Mann–Whitney U test identified variables with effect size >0.10 and p <0.05, retaining 28 radiomic features for model training (Figure 4A). Full statistical details for all variables are presented in Supplementary Material III.

**Figure 4.**
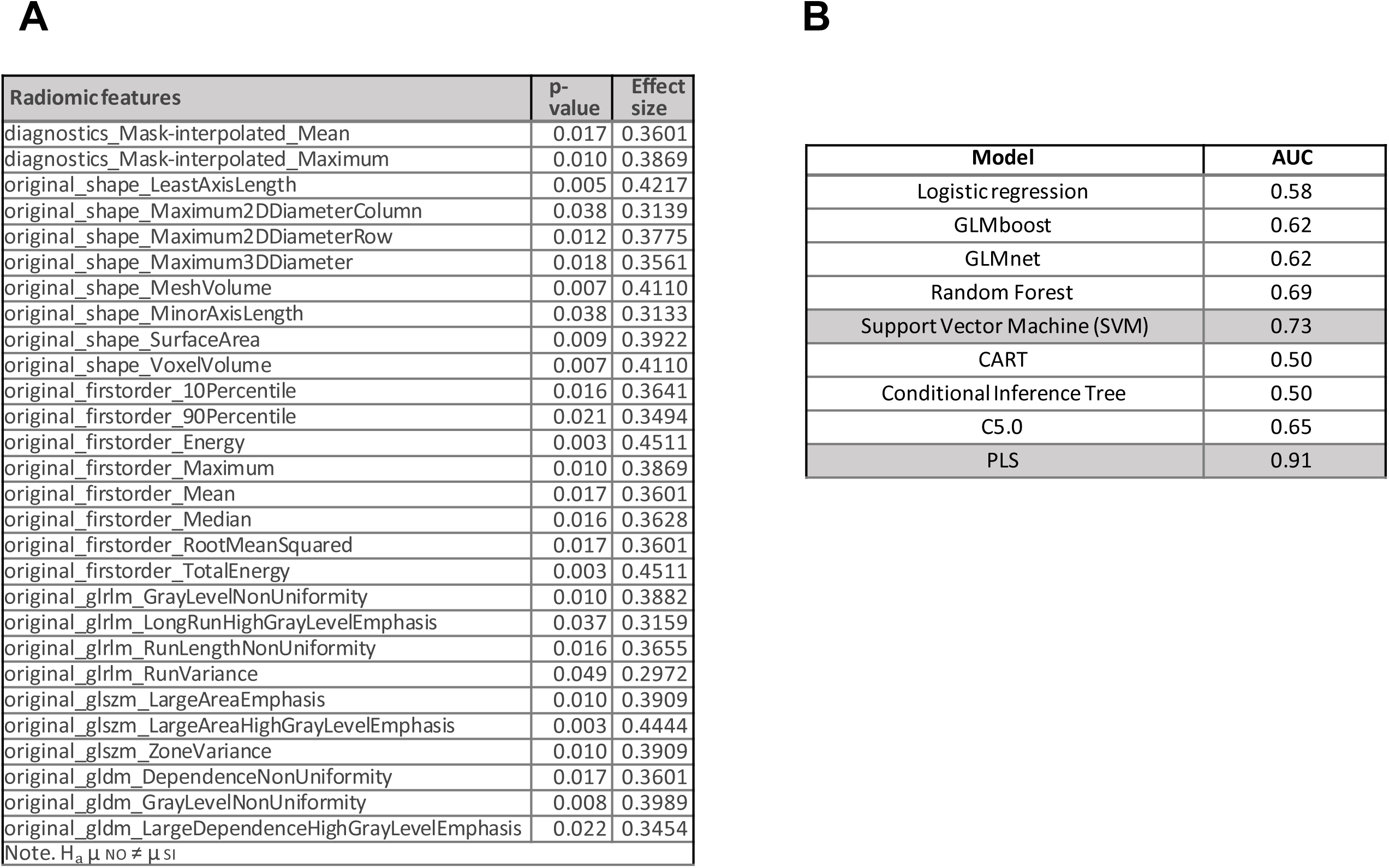
Radiomic data and model performance for the prediction of 5-year relapse in colon cancer patients. A) Radiomic features included in the 5-year relapse prediction analysis. B) AUC values of the different classification models tested for 5-year relapse prediction.

As in the previous analysis, the dataset was divided into 70% for training and 30% for testing, with 10-fold cross-validation applied. Among the models evaluated (Figure 4B), the Partial Least Squares Discriminant Analysis (PLS) and Support Vector Machine Analysis (SVM) achieved the best predictive performance, with the PLS model showing the highest AUC. Therefore, the results for the PLS and the SVM were analysed in detail.

#### 4.3.1. Partial Least Squares Discriminant Analysis (PLS)

The confusion matrices for the training and testing sets are shown in Figures 5A and 5B, respectively. The PLS model achieved a sensitivity of 0.950, effectively identifying patients who did not experience relapse within 5 years of diagnosis (Figure 5C). However, since the PLS model relies on clusters of variables, interpreting the contribution of individual predictors is challenging (Figure 5D). The ROC curve for 5-year relapse prediction showed an AUC of 0.910 (Figure 5E).

**Figure 5.**
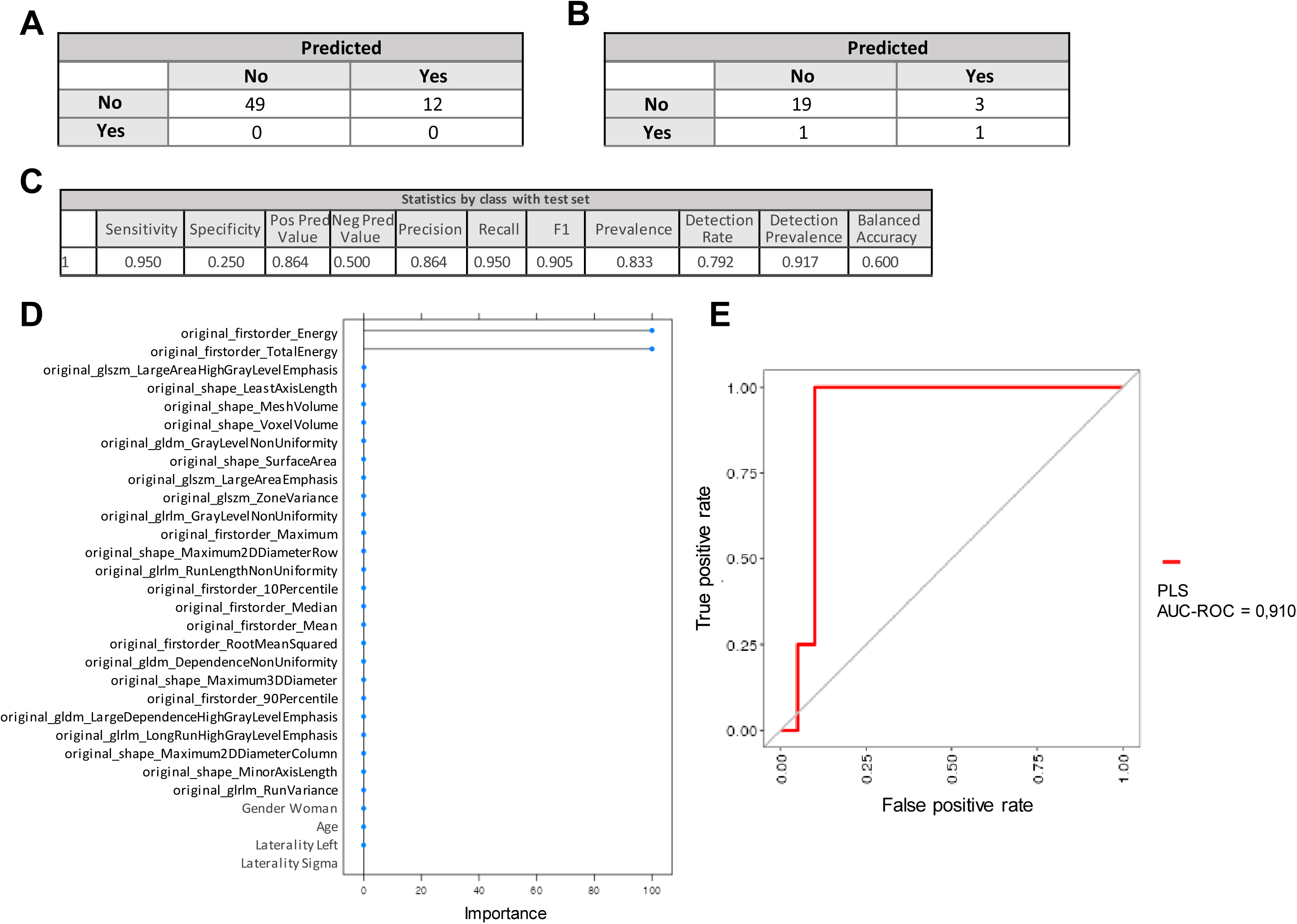
Partial Least Squares Discriminant Analysis (PLS) predicts 5-year relapse in colon cancer patients. A) Confusion matrix for the training set using the PLS model for 5-year relapse prediction. B) Confusion matrix for the test set using the PLS model for 5-year relapse prediction. C) Class statistics for the test set in the PLS model for 5-year relapse prediction. D) Variable importance in the PLS model for 5-year relapse prediction. E) ROC curve for 5-year relapse prediction using the PLS model.

#### 4.3.2. Support Vector Machine Analysis (SVM)

Given the limited interpretability of the PLS model, an SVM classifier was also implemented. Confusion matrices for the training and testing sets are shown in Figures 6A and 6B. Compared with the PLS model, the SVM exhibited major specificity and slightly lower sensitivity but still achieved robust performance, correctly identifying 90% of patients without relapse within 5 years (Figure 6C).

**Figure 6.**
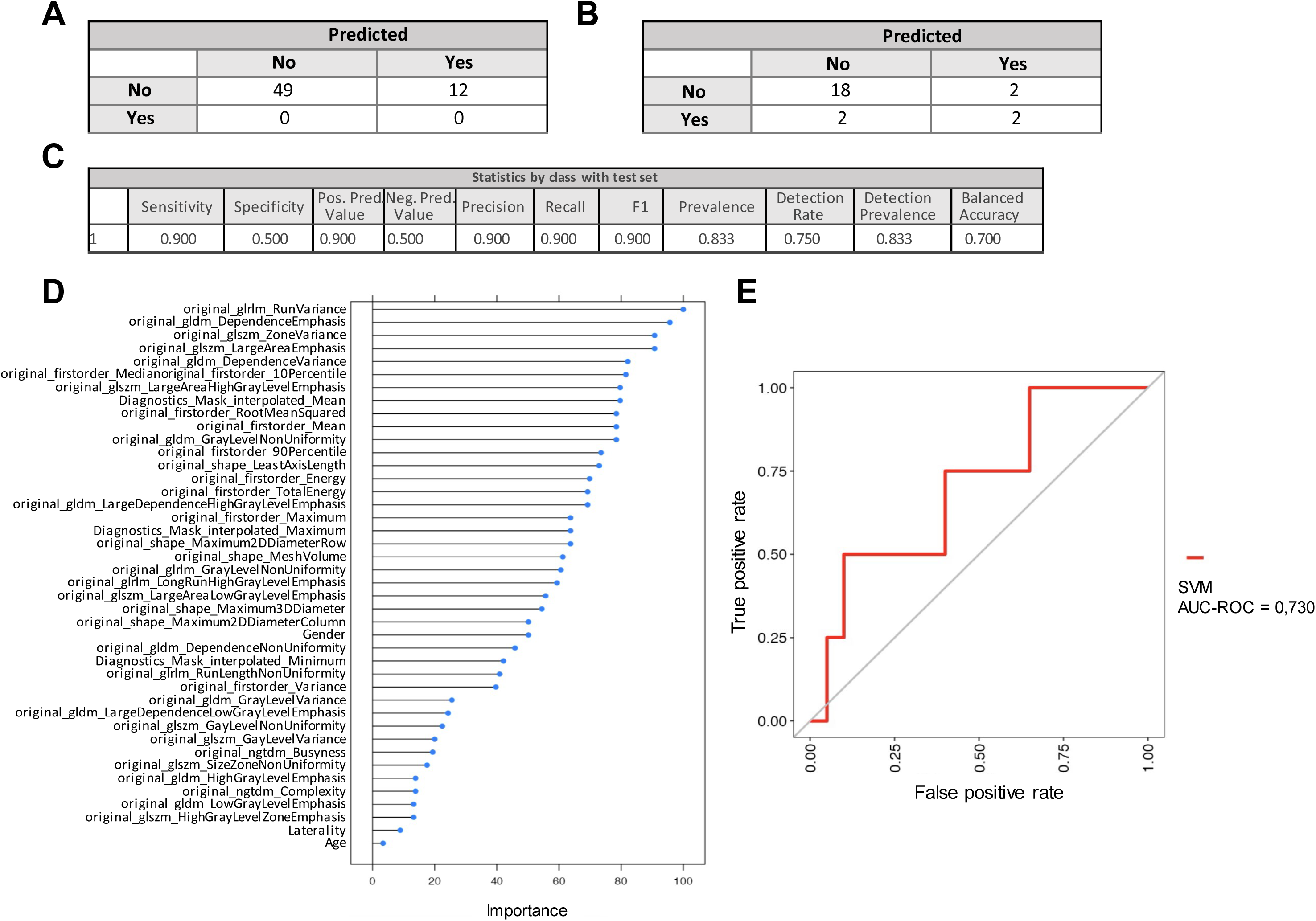
Support Vector Machine Analysis (SVM) predicts 5-year relapse in colon cancer patients. A) Confusion matrix for the training set using the SVM model for 5-year relapse prediction. B) Confusion matrix for the test set using the SVM model for 5-year relapse prediction. C) Class statistics for the test set in the SVM model for 5-year relapse prediction. D) Variable importance in the SVM model for 5-year relapse prediction. E) ROC curve for 5-year relapse prediction using the SVM model.

Unlike the PLS model, the SVM revealed that radiomic features were the most influential predictors, with the top 26 ranked variables being radiomic-derived. Among these, the Large Dependence Emphasis feature—which quantifies the distribution of large dependencies and thus reflects tissue homogeneity—was the most relevant (Figure 6D). The ROC curve for the SVM model showed an AUC of 0.730 (Figure 6E).

### 4.4. Prediction of 5-year relapse in stage II and stage III subgroups

Based on the above results, exploratory subgroup analyses were conducted to determine whether the prognostic value for predicting 5-year relapse was particularly strong within specific tumor stages (stage II and stage III). The same analytical pipeline was applied separately to each subgroup, including 40 stage II and 44 stage III patients.

#### 4.4.1. Stage II subgroup analysis

An exploratory model was developed using only stage II patients to identify those at higher risk of relapse within 5 years of diagnosis, an especially relevant question for treatment decision-making in this intermediate-risk group.

After applying variable reduction and univariate filtering (Mann–Whitney U test, effect size >0.10, p <0.05), 21 radiomic variables were retained (Figure 7A; Supplementary Material IV). Because of the smaller sample size, multicollinearity was evaluated using the Variance Inflation Factor (VIF). Variables with VIF >4 or tolerance <0.25 were considered potentially problematic (Figure 7B). Stepwise exclusion of variables with high VIF values resulted in a final model including four radiomic features with VIF <5 (Figure 7C). A binomial logistic regression model was then built without multicollinearity issues (Figure 7D).

**Figure 7.**
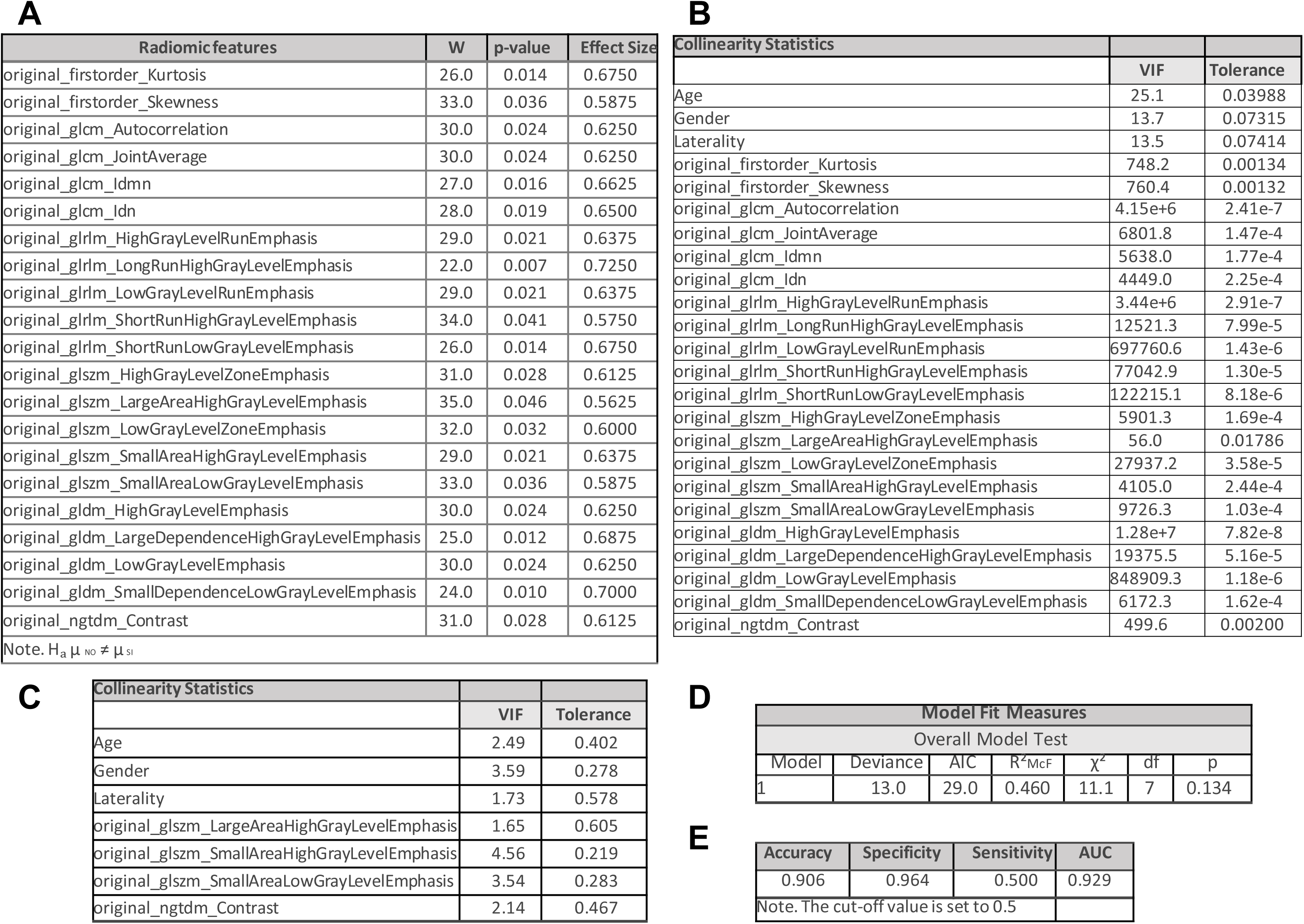
Preoperative radiomic data predict 5-year relapse in stage II CC patients. A) Radiomic features included in the analysis for 5-year relapse prediction in the stage II subgroup. B) Collinearity statistics for the exploratory analysis. C) Collinearity statistics for the final predictors. D) Model fit measures for the exploratory objective. E) Predictive performance metrics.

Although the results did not reach statistical significance due to sample size limitations, the model achieved a specificity of 0.964 and an AUC of 0.929, indicating a strong predictive capacity to identify stage II patients likely to remain relapse-free at 5 years (Figure 7E).

#### 4.4.2. Stage III subgroup analysis

Following the same analytical approach, 22 radiomic variables were retained after univariate filtering (effect size >0.10, p <0.05) (Figure 8A; Supplementary Material V). After reducing multicollinearity, seven radiomic features were included in the final model with acceptable VIF values (Figure 8B).

**Figure 8.**
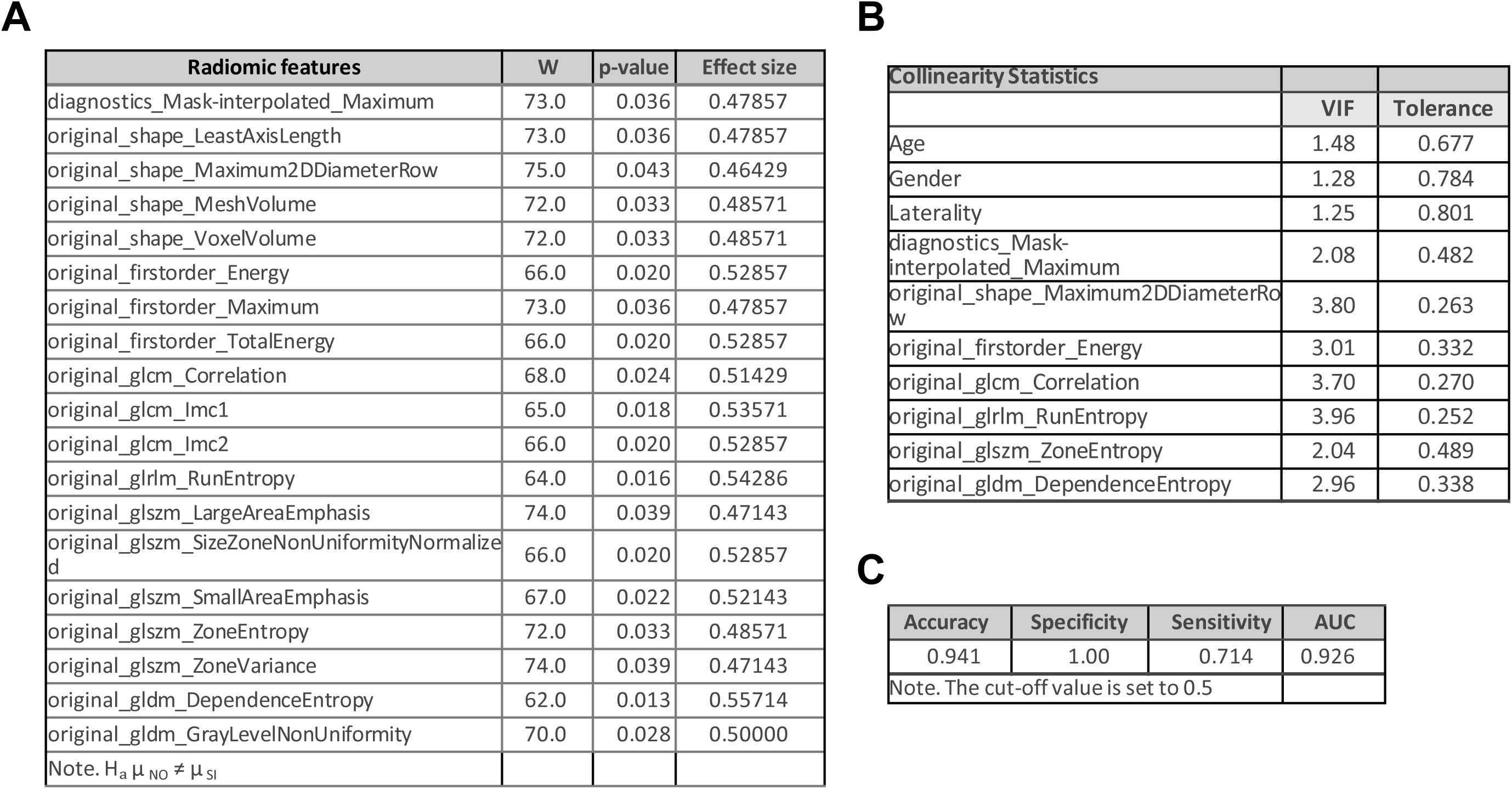
Preoperative radiomic data predict 5-year relapse in stage III CC patients. A) Radiomic features included in the analysis for 5-year relapse prediction in the stage III subgroup. B) Collinearity statistics for the final predictors. C) Predictive performance metrics.

The binomial logistic regression model, free from multicollinearity, achieved specificity and sensitivity values of 1.000 and 0.714, respectively, demonstrating strong predictive power to distinguish stage III patients who remained disease-free from those who experienced relapse within 5 years of diagnosis (Figure 8C).

## 5. DISCUSSION

In the present study, we evaluated several predictive models to discriminate between stage II and stage III CC patients using imaging biomarkers extracted from baseline preoperative CT scans and three preoperative clinical characteristics. Although the C5.0 model showed slightly higher accuracy, our results demonstrated that the GLM achieved a better balance between interpretability and performance (AUC = 0.760; sensitivity = 88.9%). In this model, clinical variables were relevant predictors, followed by four radiomic features.

We also assessed the accuracy of these models to predict the risk of disease relapse within five years after diagnosis using the preoperative radiomic features and the same three preoperative clinical characteristics. The most accurate model for predicting 5-year relapse was the PLS model (AUC = 0.910; sensitivity = 95.0%). However, the interpretability of PLS models is limited, as the method generates latent components from linear combinations of all input features, making it difficult to isolate the contribution of any single variable. Therefore, we focused on the SVM model (AUC = 0.730; sensitivity = 90.0%), where the most influential predictors were radiomic texture features. Notably, more homogeneous textures were associated with the absence of relapse during follow-up, suggesting a link between image heterogeneity and tumor aggressiveness. All these findings highlight the potential of radiomic features as non-invasive imaging biomarkers that could support clinical decision-making and improve patient management in CC.

Previous studies have explored the role of radiomics in CC. Some have proposed radiomic signatures for recurrence and survival prediction (Caruso et al., 2022) (Dai et al., 2020) (JM et al., 2021) (Li et al., 2019), while others focused on tumor characterization and local staging (Li et al., 2019) (Zerunian et al., 2024). In addition, radiomics has been applied to predict molecular biomarkers such as microsatellite instability (MSI) (Golia Pernicka et al., 2019), as well as histopathological prognostic factors including perineural invasion (Guo et al., 2024). However, the number of studies specifically addressing CC remains limited. Most radiomic analyses have been performed on broader colorectal cancer cohorts, where colon and rectal tumors are jointly evaluated (Rompianesi et al., 2022) (Lv et al., 2022) (Badic et al., 2019) (Li et al., 2022) (M et al., 2021) (Rocca et al., 2021). Given that the colon and rectum differ anatomically, vascularly, and neurologically—with distinct metastatic spread patterns (Riihimaki et al., 2016)—they should be analyzed independently. Accordingly, focusing exclusively on CC, as done in the present study, provides more biologically and clinically relevant insights and enables more accurate patient stratification for tailored therapeutic strategies.

Approximately 25–30% of CC patients develop liver metastases (Engstrand et al., 2018), the leading cause of CC-related death. Stage III CC patients are known to have a high recurrence risk, though a small proportion of stage II patients also experience relapse (Shi et al., 2013). Adjuvant chemotherapy clearly improves outcomes in stage III disease, increasing 5-year DFS from 49.0% to 63.6% (Siegel et al., 2019), whereas its benefit in stage II remains controversial(Böckelman et al., 2015). Prognosis in both stage II and stage III is influenced by multiple factors, including tumor (pT) and nodal (pN) stage, histologic grade, number of lymph nodes examined, vascular invasion, mismatch repair (MMR) status, and surgical urgency (Böckelman et al., 2015). Among stage II patients, DFS may decrease from 91% to 75% depending on these risk factors (Gertler et al., 2009). Interestingly, certain stage III patients may have relatively low recurrence risk and favorable outcomes, even with shorter chemotherapy regimens that reduce cumulative neurotoxicity (Grothey et al., 2018). Consequently, some stage III patients without additional risk factors may have a better prognosis than high-risk stage II patients. However, clinic-pathological markers remain insufficient to accurately stratify risk within these stages, leaving a critical gap in identifying the subset of high-risk stage II patients who may benefit from chemotherapy and the low-risk stage III patients who might be candidates for de-escalation.

As an exploratory objective, we separately analyzed the prediction of 5-year relapse in stage II and stage III subgroups. Interestingly, the models stratified by stage achieved excellent performance, with an AUC = 0.929 for stage II (specificity = 96.4%) and an AUC = 0.926 for stage III (specificity = 100%). Nevertheless, this remains a pilot study, and the small cohort sizes preclude definitive conclusions. Larger, multicenter studies are needed to validate these findings.

This study has several limitations. First, it is a single-center retrospective analysis, which may introduce selection bias. Future work should include prospective, multicenter validation to enhance generalizability. Second, the limited sample size constrains model robustness and should be expanded to further refine predictive performance.

In conclusion, we developed CT-based radiomic models capable of accurately distinguishing between stage II and stage III CC patients and predicting 5-year relapse after diagnosis. Radiomic features contributed significantly to model performance, particularly those describing tumor texture heterogeneity. Our exploratory findings also suggest that incorporating radiomic data into initial staging could improve patient risk stratification. Therefore, these non-invasive radiomic models hold promise as novel tools for personalized management of CC patients, potentially guiding therapeutic decisions and improving patient quality of life.

## INFORMED CONSENT STATEMENT

Informed consent was obtained from all subjects involved in the study.

## Data Availability

All data produced in the present study are available upon reasonable request to the authors

## ACKNOWLEDGMENTS

ChatGPT was used for English text editing. We are grateful to lab members for help and advice throughout this research.

